# An Open-Source Generalizable Deep Learning Framework for Automated Corneal Segmentation in Anterior Segment Optical Coherence Tomography Imaging

**DOI:** 10.1101/2025.06.18.25329856

**Authors:** Lynn Kandakji, Siyin Liu, Shafi Balal, Ismail Moghul, Bruce Allan, Stephen Tuft, Daniel Gore, Nikolas Pontikos

## Abstract

**Purpose:** To develop a deep learning model – Cornea nnU-Net Extractor (CUNEX) – for full-thickness corneal segmentation of anterior segment optical coherence tomography (AS-OCT) images and evaluate its utility in artificial intelligence (AI) research.

**Methods:** We trained and evaluated CUNEX using nnU-Net on 600 AS-OCT images (CSO MS-39) from 300 patients: 100 normal, 100 keratoconus (KC), and 100 Fuchs endothelial corneal dystrophy (FECD) eyes. To assess generalizability, we externally validated CUNEX on 1,168 AS-OCT images from an infectious keratitis dataset acquired from a different device (Casia SS-1000). We benchmarked CUNEX against two recent models, CorneaNet and ScLNet. We then applied CUNEX to our dataset of 194,599 scans from 37,499 patients as preprocessing for a classification model evaluating whether segmentation improves AI prediction, including age, sex, and disease staging (KC and FECD).

**Results:** CUNEX achieved Dice similarity coefficient (DSC) and intersection over union (IoU) scores ranging from 94–95% and 90–99%, respectively, across healthy, KC, and FECD eyes. This was similar to ScLNet (within 3%) but better than CorneaNet (8–35% lower). On external validation, CUNEX maintained high performance (DSC 83%; IoU 71%) while ScLNet (DSC 14%; IoU 8%) and CorneaNet (DSC 16%; IoU 9%) failed to generalize. Unexpectedly, segmentation minimally impacted classification accuracy except for sex prediction, where accuracy dropped from 81 to 68%, suggesting sex-related features may lie outside the cornea.

**Conclusion:** CUNEX delivers the first open-source generalizable corneal segmentation model using the latest framework, supporting its use in clinical analysis and AI workflows across diseases and imaging platforms. It is available at https://github.com/lkandakji/CUNEX.

## Introduction

Anterior segment optical coherence tomography (AS-OCT) provides high-resolution radial cross-sections of the cornea that can support the diagnosis and management of corneal diseases such as keratoconus (KC),^1, 2^ Fuchs endothelial corneal dystrophy (FECD),^3^ and infectious keratitis (IK).^4^ However, interpreting these images when there is early disease requires a high level of expertise. Deep learning (DL), particularly convolutional neural networks (CNNs), has advanced image analysis,^5^ often outperforming clinician-reviewed scans or pre-selected metrics such as pachymetry, curvature, or symmetry indices.^6, 7^ However, their diagnostic performance may depend on image quality and/or limiting the analysis on the most relevant anatomical regions.^8,9^

To date, most DL image analysis models have been trained on data from Placido disk or Scheimpflug-derived topographic maps.^10^ Despite growing interest in AS-OCT, cross-sectional images, sometimes referred to as B-scans, are still rarely reported in DL classification studies. In other imaging domains, including magnetic resonance imaging (MRI),^11^ computed tomography (CT),^12^ fundus photography,^13, 14^ and retinal OCT, ^15, 16^ segmentation has improved performance and interpretability. Segmentation delineates a pathological region or anatomical structure, enabling a structure-specific input for further analysis, but its role as part of the model workflow for the diagnosis of corneal disease remains untested.

AS-OCT classification workflows generally follow two approaches: (1) Measurement-based classification, where biometric parameters are extracted from the AS-OCT B-scan and these numeric values are used as the input for a classifier; ^17–19^ or (2) End-to-end DL, where a CNN analyzes the AS-OCT image output from either a cross-section or derived heatmap and predicts a condition.^20–22^ Segmentation may enhance either approach by isolating the most diagnostically relevant structures and reducing noise from surrounding anatomy.

We developed a novel cornea nnU-Net extractor (CUNEX), a generalizable DL framework that performs full thickness limbus-to-limbus corneal segmentation from AS-OCT cross-sectional images. Our goal was to create an accurate and adaptable segmentation tool to support a wide range of AS-OCT research and clinical applications. To inform future modeling priorities for AS-OCT analysis, we also investigate the role of corneal segmentation in DL classification of AS-OCTs and determine when anatomical precision meaningfully contributes to model utility.

## Materials and Methods

### Dataset

The study received ethical approval from Moorfields Eye Hospital (MEH) NHS Foundation Trust (references CA17/CED/03 and 22/PR/0249). The study adhered to the principles outlined in the Declaration of Helsinki.

The Moorfields Eye Hospital Corneal dataset included 194,599 AS-OCT scans (MS-39, Costruzione Strumenti Oftalmici, Florence, Italy) from 37,499 patients. Each scan consists of 25 radial cross-sections with a resolution of 900×1800 pixels per image. To train CUNEX, we defined three corneal classes: 100 unique normal eyes, 100 that had KC, and 100 with FECD. For the classification CNN, we selected a separate set of 600 unique healthy eyes, 480 with KC, and 400 with FECD. Clinical diagnoses were confirmed by a fellowship-trained corneal specialist following review of the AS-OCT images and electronic health records. The normal eyes were from patients reviewed before laser vision correction (LVC) with no ocular pathology apart from myopia.

Each model development dataset was split into training (70%), test (15%), and validation (15%), stratified by class (**Figure 1**). Model performance was evaluated on the validation set. All training was performed on an NVIDIA GeForce RTX 3090 GPU.

**Figure 1.**
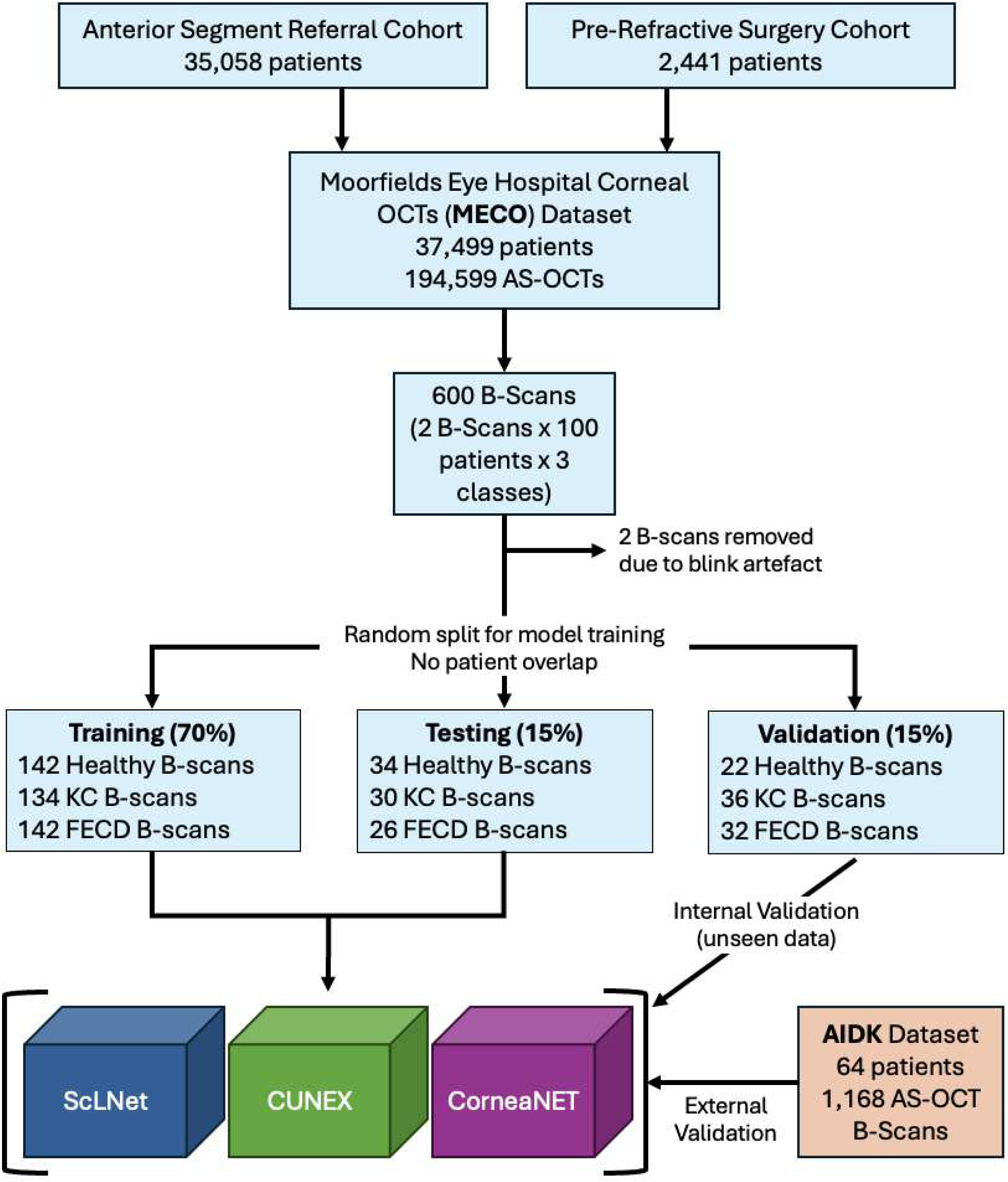
Dataset selection and partitioning workflow.

To assess CUNEX generalizability on out-of-distribution data - i.e., images that differ in pathology, resolution, and device from those used in training - and simulate a real-world clinical deployment we performed external validation without additional retraining using an independent dataset of eyes with infectious keratitis (IK).^23^ This dataset comprises 1,168 AS-OCT images with resolution (1723 × 1000 pixels) from 64 patients with IK from a swept-source Casia SS-1000 OCT system (Tomey Corporation, Japan).

### Manual Annotation

To create the labels used to train CUNEX, AS-OCT images were manually annotated (LK) on the Moorfields Grading Platform (grading.readingcentre.org). In this context, labels refer a single manually drawn region covering the whole of the imaged section of the cornea, which serves as the reference (ground-truth) for training the segmentation model. From each 25-scan volume, a transverse and sagittal view was selected to delineate the whole cornea from limbus to limbus (**Supplemental Digital Content Figure 1**). To assess reproducibility, a subset of 50 scans was independently annotated by a second grader (RL) and reviewed by a fellowship-trained corneal specialist (ST).

### Segmentation Model Development and Evaluation

We developed CUNEX using the nnU-Net framework. For comparison, we used two other U-Net models, CorneaNet^24^ and ScLNet.^25^ As our image dimensions were incompatible with CorneaNet, we retrained its architecture on our internal dataset. A pre-trained ScLNet model could not be accessed, so we also retrained its architecture on our internal dataset. We retained the original training hyperparameters and number of epochs for both CorneaNet and ScLNet, as specified in their source codes. The architectural and pre-processing differences between CUNEX and previous U-Net implementations are detailed in **Supplemental Digital Content Table 1**.

We assessed segmentation performance against manual annotation using three metrics: the Dice Similarity Coefficient (DSC), intersection over union (IoU), and the Hausdorff Distance (HD). DSC and IoU measure the overlap between the predicted and manual segmentation masks, while HD assesses the boundary accuracy. To assess segmentation consistency, each corneal image was vertically divided into three equal segments and DSC, IoU, and HD were calculated for the central and peripheral zones.

The segmentation model generalizability was assessed using two validation sets. First, an internal hold-out set from the same device (MS-39). Then, the trained model weights were applied without re-training or fine-tuning to an external dataset (AIDK). Segmentation ground truth for the AIDK set was provided by the authors as polygon-based labels (LabelMe JSON format). We then quantified segmentation accuracy by comparing the CUNEX-predicted masks to the manual reference annotations.

### Classification Model Development and Evaluation

We conducted four binary classification experiments to evaluate the impact of segmenting the cornea with CUNEX on classification performance. Two experiments focused on discriminating between early- vs end-stage KC and FECD and another two on discriminating age (≤30 or >30) and sex (male or female). Age and sex predictions used only healthy eyes to avoid confounding by disease-related patterns, as KC and FECD affect younger^24^ and older^25^ individuals, respectively. We empirically selected the 30-year age threshold based on the distribution of ages in the healthy cohort (mean 30 ± 8 years, median 29, range 16–60) to yield a balanced split. To address class imbalance in the training data for early vs end stage KC and FECD, we used weighted sampling so that the CNN saw a balanced number of examples from each class during learning.

Each experiment was conducted twice using two input types: (1) the original AS-OCT cross-sectional images (classification-only), and (2) the same images masked to show only the cornea (segmentation-classification). In the second approach, segmentation masks generated by CUNEX were applied to remove non-corneal regions. The CNN architecture was kept consistent across all experiments and was based on lightweight classification models such as MobileNet^26^ and Xception^27^ (**Supplemental Digital Content Table 2**). All models were trained for 10 epochs using the Adam optimizer and binary cross-entropy loss. Rather than optimizing the network architecture itself, the aim of this study was to evaluate whether providing an explicit anatomical focus – the cornea – improves the CNN’s ability to correctly classify the input.

Model performance for each binary classification task was evaluated on a held-out validation set using five standard metrics: accuracy, sensitivity (recall), specificity, F1 score, and Matthews correlation coefficient (MCC). We used F1 score and MCC over receiver operating characteristic curves and area under the curve, to reflect clinical use of fixed decision thresholds and performance under class imbalance. All metrics were computed separately for each of the four classification tasks. To compare the two input conditions, we used McNemar’s test with statistical significance set at *p* < 0.05.

## Results

### Segmentation Training

One volume from the healthy group was excluded from the segmentation dataset due to motion artifact caused by blinking. Inter-grader agreement of manual annotations (n=98) was high with an average DSC of 98%, IoU of 84%, and ICC for mask area of 0.83 (r=0.83, p<0.001). Most boundary differences were <60 pixels with a mean HD of 130.9 ± 97.4 (**Supplemental Digital Content Table 3**). CUNEX and ScLNet reached 96% DSC after 205 and 100 training epochs, respectively, while CorneaNet achieved 91% after 600 epochs.

### Quantitative Validation

On the internal validation set, CUNEX consistently outperformed both CorneaNet and ScLNet, particularly in KC and FECD cases (**Table 1**). While CUNEX and ScLNet achieved similar DSC and IoU scores, CUNEX showed more localized boundary errors, reflected in slightly higher HD values. In contrast, CorneaNet exhibited substantially lower overlap metrics, especially in KC, and produced the largest and most spatially diffuse boundary errors, indicating poor generalizability to atypical corneal shapes. Segmentation accuracy was highest in the central cornea across all models, likely due to clearer anatomical contrast and fewer non-corneal structures. However, HD values were paradoxically greater in the central zone, suggesting the presence of small but sharp deviations in predicted boundaries near the apex. On the external AIDK dataset, CUNEX maintained strong performance with DSC and IoU scores above 80 and 70%, respectively. In contrast, CorneaNet and ScLNet failed to generalize, with DSC and IoU scores less than 16 and 10%, respectively.

**Table 1.**
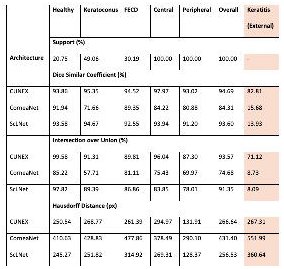
Performance of various models on validation data is presented. Support represents the proportion of images each class occupies within the dataset. Both the DSC and IOU scores for each of the three image classes are detailed, alongside comparisons between the central (inner third) and peripheral regions (outer thirds) of the OCT image.

### Qualitative Validation

Visual inspection confirmed that CUNEX consistently isolated the whole corneal structure across diverse presentations while excluding surrounding features such as the limbus, iris, lens, and sclera (**Figure 2**). It handled common imaging artifacts with minimal degradation, including central lines, speckle noise, and low SNR regions. ScLNet demonstrated similar performance to CUNEX but with slightly coarser boundary definitions and localized discontinuities, suggesting reduced subpixel precision at the epithelial and endothelial margins. In contrast, CorneaNet exhibited frequent segmentation failures, including incomplete boundaries, discontinuous regions, and false positives in adjacent structures, particularly in KC and FECD images.

**Figure 2.**
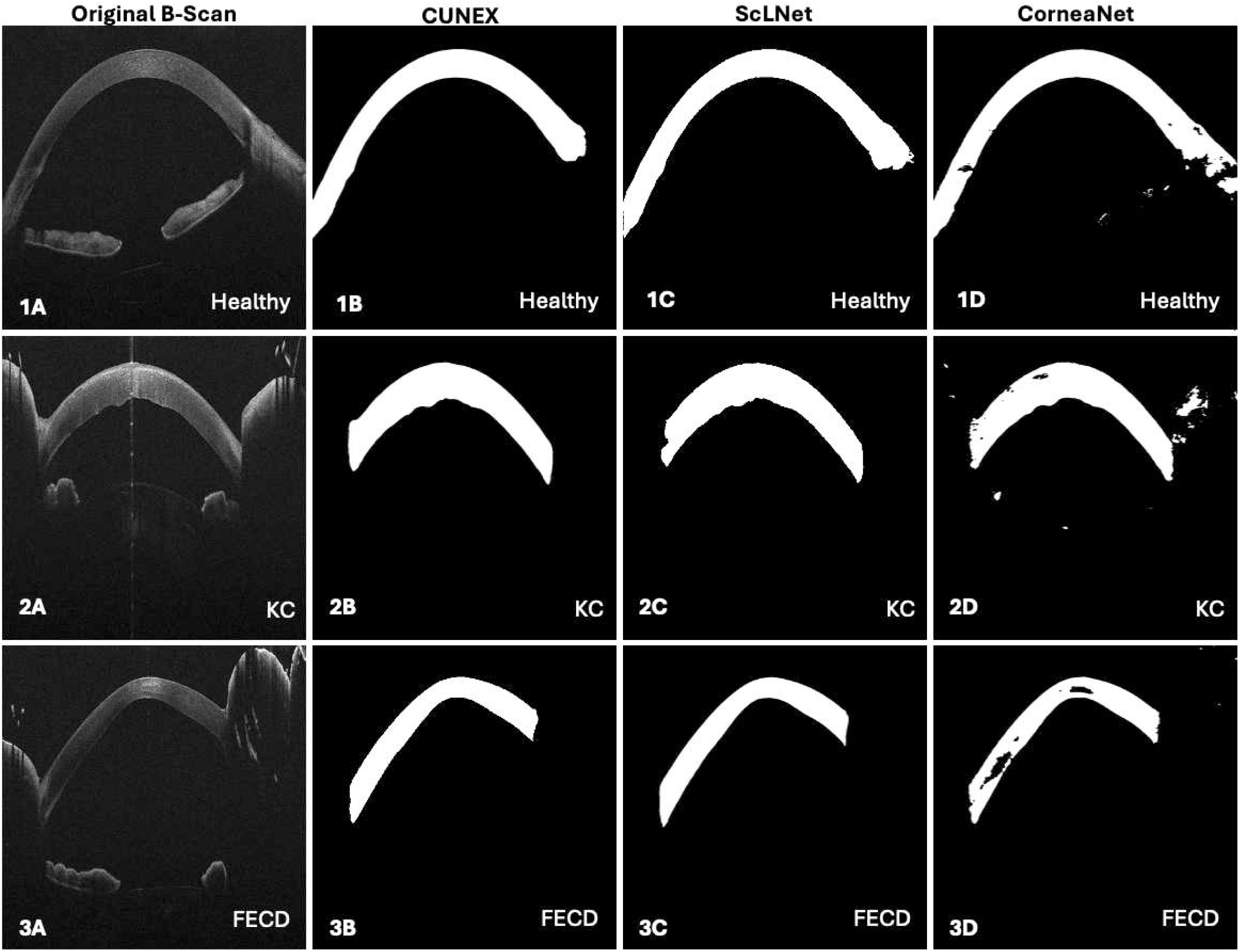
Variability in the performance of three segmentation networks: CUNEX, CorneaNet, and ScLNet, in segmenting the cornea across healthy, keratoconus (KC) and Fuchs endothelial corneal dystrophy (FECD). Each row shows a representative cross-section AS-OCT image from a healthy eye (row 1), KC eye (row 2), and eye with FECD (row 3). Column show the original B-scan (A), and corresponding corneal segmentation mask predicted by CUNEX (B), ScLNet (C), and CorneaNet (D).

On the external AIDK dataset, segmentation performance diverged significantly between the three models (**Figure 3**). CUNEX demonstrates strong generalizability, producing anatomically accurate outputs across all cases. However, prominent central artifact lines near the visual axis in AIDK images occasionally caused central overcuts in CUNEX’s outputs and likely contributed to its slight drop in validation accuracy. In contrast, ScLNet produced large false positives and anatomically implausible extensions beyond the corneal boundaries, while CorneaNet captured only partial and disjointed anterior and posterior corneal boundaries.

**Figure 3.**
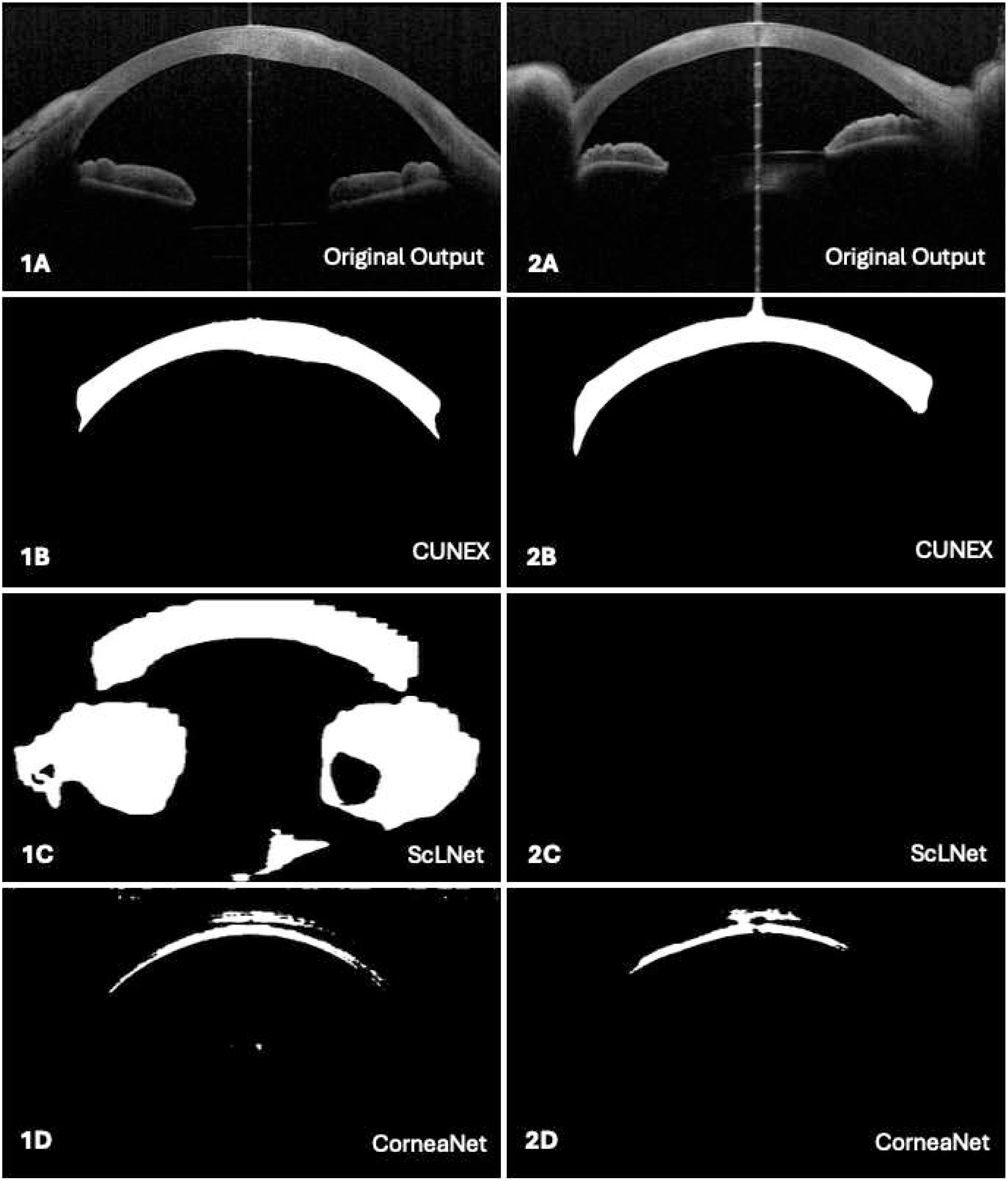
External segmentation performance on keratitis AS-OCT images (AIDK dataset) acquired using the Casia SS-1000 swept-source device. Row A displays the original cross-section image, row B the CorneaNet prediction, row C the ScLNet prediction and row D the CUNEX prediction.

### Comparative Evaluation of Classification Performance with and without Segmentation

Segmentation did not improve discriminative ability in any of the four classification tasks. Performance metrics were consistently, but marginally, lower with segmentation-assisted inputs compared to the original image (**Table 2**). Most prediction differences involved a single false positive or negative (**Figure 4**), suggesting normal variability in training. Segmentation slightly reduced specificity in KC staging and improved it in FECD staging, though neither change was statistically significant. Sex classification showed a significant decrease in performance with the segmented input, which may indicate that sexually dimorphic features are located outside the cornea and are partially suppressed with segmentation.

**Figure 4.**
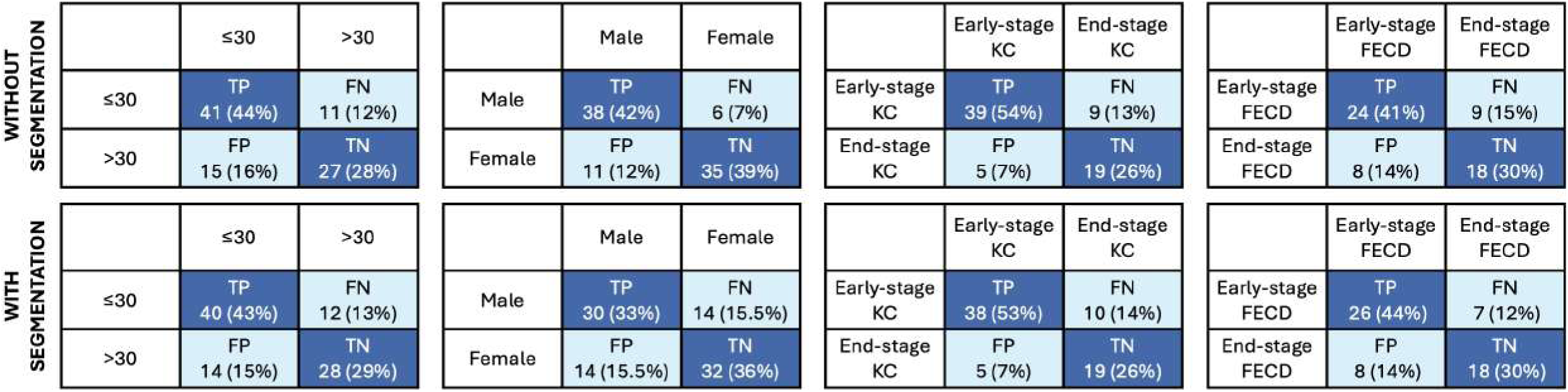
Confusion matrices for binary classification performance of deep learning models trained to predict age group (≤30 vs >30), sex (male vs female), and disease stage (early vs end-stage keratoconus or FECD) in AS-OCT images. Performance is reported on validation data, comparing models trained without (top row) and with (bottom row) corneal segmentation applied to the input image. True labels were derived from clinical records. Each cell shows absolute counts with percentage of total. TP=true positive; TN=true negative; FP=false positive; FN=false negative; KC=keratoconus; FECD=Fuchs’ endothelial corneal dystrophy.

**Table 2.**
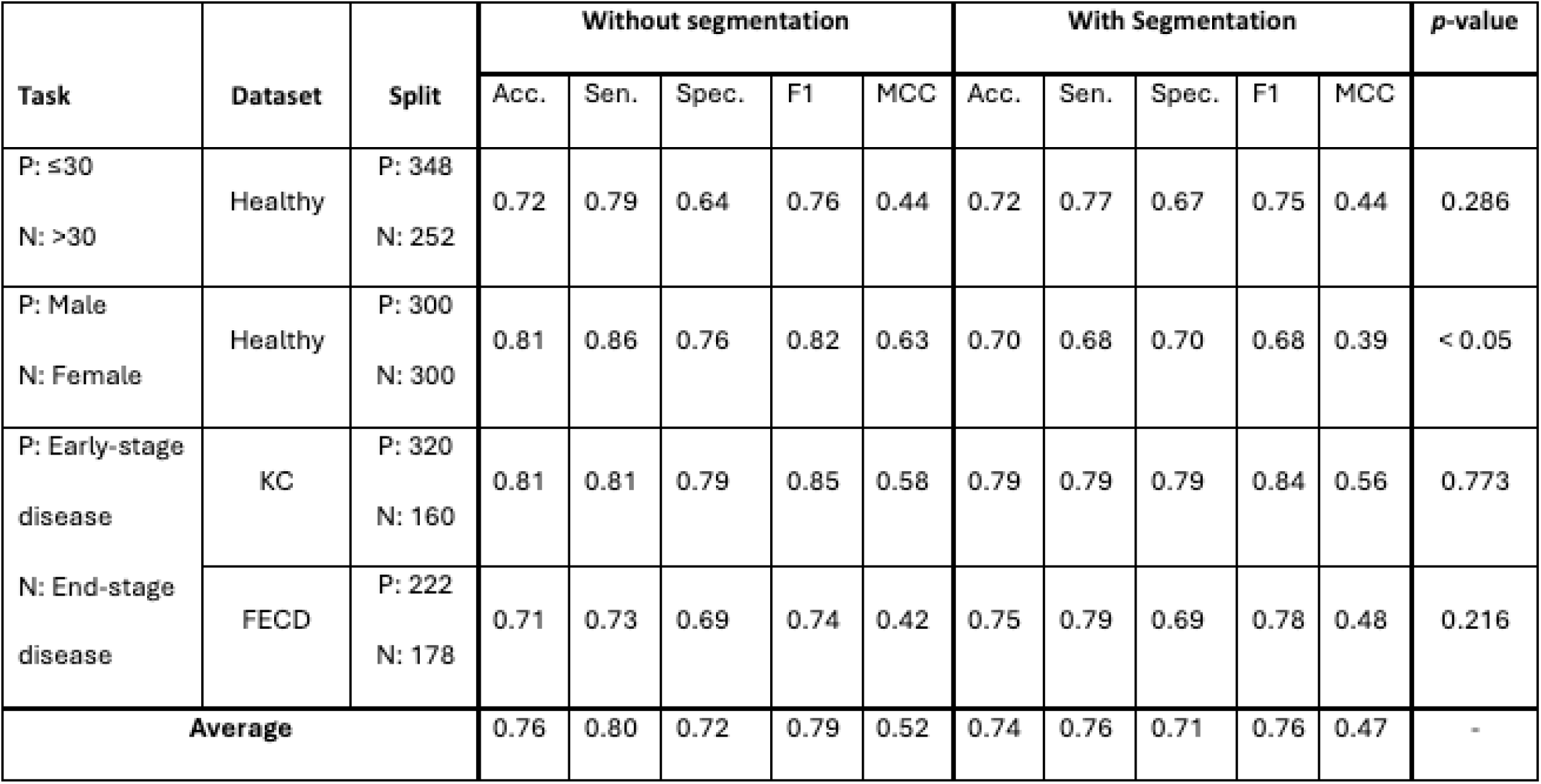
Performance of the classification model in various binary tasks on the testing set with and without segmentation of images prior to training. P=Positive predictor, N=Negative predictor, Acc=Accuracy, Sen=Sensitivity, Spec=Specificity, MCC=Matthews correlation coefficient.

## Discussion

This paper introduces CUNEX, the first externally validated DL model for full-thickness segmentation in AS-OCT cross-sectional images. We trained CUNEX using a standardized, data-driven framework (nnU-Net), avoiding manual configuration and investigator bias. CUNEX achieved high segmentation accuracy cross healthy, KC, and FECD groups. We compared CUNEX to two open-source AS-OCT segmentation models, ScLNet and CorneaNet, retrained both on our internal dataset, and tested all three models on a held-out internal validation set as well as an external IK dataset from a different AS-OCT device. We also assessed the impact of segmentation on disease staging and age and sex discrimination by training a CNN classifier with and without CUNEX-generated segmentation masks.

ScLNet and CorneaNet are established AS-OCT segmentation models. ScLNet employs multiscale contextual encoding and edge-aware supervision, whereas CorneaNet adopts a variant of the classic U-Net structure.^28^ Both were reported to achieve high accuracy (DSC, 99% for both; IoU 98% and 97%,respectively) and were validated on healthy and KC eyes. CorNet,^29^ another AS-OCT segmentation model, was excluded from our comparison as it is not open source and thus not reproducible. AS-OCT scans capture multiple interfaces and anatomy in one image, including the cornea, iris, lens, sclera, the eyelid and lashes, making full corneal segmentation complex. Prior studies have trained segmentation models on cropped portions of the image, such as the anterior chamber angle or central cornea.^30–32^

On the internal validation set, CUNEX performed similarly to ScLNet, with DSC and IoU score differences under 3%, and consistently outperformed CorneaNet across all classes, particularly KC. This may be because CorneaNet was trained on images from a custom-built ultrahigh-resolution spectral-domain OCT (UHR-SD-OCT) system^33^ not available for clinical use.^33^ This highlights a limitation of generalizability of models developed on single device datasets.

CUNEX delivers robust corneal segmentation across heterogenous AS-OCT inputs, such as variation in corneal morphologies, devices, image quality and resolutions, without retraining. On an external dataset of IK images with a different resolution acquired using a different AS-OCT device,^34^ CUNEX still produced reliable corneal segmentation, whereas ScLNet and CorneaNet did not. This capability addresses domain shift, a persistent challenge in real-world DL deployment where a model trained on one dataset performs poorly on data from another device.^35, 36^ CUNEX’s data-driven configuration enables it to learn transferable anatomical representations rather than overfitting a specific image input, make it well-suited for multi-center studies and explainable AI.

It has not been tested whether integrating segmentation improves DL classification performance for anterior segment diseases. In our experiments, corneal segmentation did not improve classification accuracy, suggesting that the CNN is able to identify the relevant features within the original input image and that predictions were primarily driven by corneal anatomy. However, the sex classification performance decreased significantly when segmentation was applied to the input image, reducing accuracy to near chance levels. This implies that the anatomical features used to infer sex in AS-OCT images may lie outside the cornea. Previous studies using AS-OCT have linked sex differences to corneal shape, anterior chamber conformation, and lens position.^37^ Rather than simply a means to potentially boost performance, segmentation may serve as a tool to probe which regions meaningfully influence a given classification task.

Ultimately, CUNEX is the only externally validated open-source model for whole corneal segmentation in AS-OCT. It is based on the latest architecture for segmentation, nnU-Net, and is generalizable, which means new users will not have to retrain CUNEX to run inference on their dataset, compared to prior models which are not as easily reproducible. This could support broader AS-OCT research and use in multi-center studies and low-resource settings, where imaging protocols and resolution may vary. Besides classification models, it can be used for a range of additional applications such as biomechanical modelling that relies on precise corneal boundaries and surgical planning tools requiring accurate anatomical mapping.

There are some limitations. Both ScLNet and CorneaNet were originally developed for more complex segmentation tasks, including multi-class labeling of individual corneal layers or differentiation of the cornea from scleral lenses. For the purposes of this study, we adapted both models to perform a simplified task—segmenting only the whole cornea—to allow direct comparison with CUNEX. Segmentation also adds complexity, requiring expert annotations and processing time, which must be balanced against potential gains in predictive accuracy. Some studies suggest that when training datasets are large – on the order of thousands of images – and the classification involves easily distinguishable classes, such as healthy vs diseased eyes, CNNs can learn relevant features without requiring segmentation.^38^ These factors may explain why segmentation has not been widely adopted as a pre-processing step, but could become increasingly helpful as models evolve towards detecting subtler features, such as those seen in early-stage disease.

As experimental adoption of volumetric AS-OCT increases, following trends in MRI imaging,^11^ future work could explore 3D segmentation of the full cornea to improve spatial continuity and enable more precise mapping of focal pathology or surgical planning. A 2D model was used in this study to reflect the current standard of AS-OCT research and maximize computational efficiency, but the nnU-Net framework can be readily extended to 3D volumetric data. Moreover, although AS-OCT provides detailed visualization of anterior segment anatomy, its integration into interpretable AI frameworks remains unexplored. In this context, CUNEX lays the groundwork the development of explainable AI tools in clinical practice.

## Data Availability

All data produced in the present study are available upon reasonable request to the authors.

https://github.com/lkandakji/CUNEX

## Model Availability

The source code and model weights of CUNEX have been made publicly available at https://github.com/lkandakji/CUNEX/.

## Legend for Supplemental Digital Content

Supplemental Digital Content Figure 1.

Title: Representative manually annotated AS-OCT B-scans.

Description: Example annotated transverse (A) and sagittal (B) anterior segment optical coherence tomography B-scans, showing manual segmentation of the full-thickness cornea (highlighted in yellow). The cornea is segmented from limbus to limbus, capturing anterior and posterior boundaries while avoiding the sclera.

Supplemental Digital Content Table 1.

Title: Comparison of segmentation model architectures and configurations.

Description: Comparison between standard U-Net, CorneaNet, CorNet, ScLNet, and CUNEX. CUNEX is generalizable to different input sizes and is optimized for AS-OCT images by preserving image resolution, adjusting network depth.

Supplemental Digital Content Table 2.

Title: Classification CNN architecture parameters.

Description: Layer-wise configuration of the CNN used for downstream classification.

Supplemental Digital Content Table 3.

Title: Inter-grader agreement statistics for manual annotations.

Description: Summary statistics of inter-grader segmentation agreement across 98 AS-OCT images. Reported metrics include Dice similarity coefficient (DSC), Intersection over Union (IoU), and Hausdorff distance, as well as the annotated mask area in pixels for each grader (L.K. and R.L.). All values are presented as mean, standard deviation, minimum, and maximum.

Supplemental Digital Content Figure 2.

Title: CUNEX training performance.

Description: CUNEX loss and Dice similarity coefficient (DSC) graph for fold 1, using both Dice loss and cross-entropy loss as the loss function (red and blue curves) and DSC as the accuracy metric (green curves). Loss curves for both models initially demonstrated rapid decreases, indicating rapid early learning, and subsequently stabilized, reflecting incremental model improvements with occasional spikes due to challenging data batches.

